## Supplementary Appendix for "Study Protocol: Development and Retrospective Validation of an Artificial Intelligence System for Diagnostic Assessment of Prostate Biopsies"

#### Version 1.0

#### Supplementary Appendix

Nita Mulliqi[1], Anders Blilie[2,3], Xiaoyi Ji[1], Kelvin Szolnoky[1], Henrik Olsson[1], Matteo Titus[1], Geraldine Martinez Gonzalez[1], Sol Erika Boman[1,4], Masi Valkonen[5], Einar Gudlaugsson[2], Svein R. Kjosavik[3,6], José Asenjo[7], Marcello Gambacorta[8], Paolo Libretti[8], Marcin Braun[9], Radzislaw Kordek[9], Roman Łowicki[10], Kristina Hotakainen[11,12], Päivi Väre[13], Bodil Ginnerup Pedersen[14,15], Karina Dalsgaard Sørensen[15,16], Benedicte Parm Ulhøi[17], Mattias Rantalainen[1], Pekka Ruusuvuori[4,18], Brett Delahunt[19], Hemamali Samaratunga[20], Toyonori Tsuzuki[21], Emilius A.M. Janssen[2,22], Lars Egevad[23], Kimmo Kartasalo[1], Martin Eklund[1]

1. Department of Medical Epidemiology and Biostatistics, Karolinska Institutet, Stockholm, Sweden
2. Department of Pathology, Stavanger University Hospital, Stavanger, Norway
3. Faculty of Health Sciences, University of Stavanger, Stavanger, Norway
4. Department of Molecular Medicine and Surgery, Karolinska Institutet, Stockholm, Sweden
5. Institute of Biomedicine, University of Turku, Turku, Finland
6. The General Practice and Care Coordination Research Group, Stavanger University Hospital, Norway
7. Department of Pathology, Synlab, Madrid, Spain
8. Department of Pathology, Synlab, Brescia, Italy
9. Department of Pathology, Chair of Oncology, Medical University of Lodz, Lodz, Poland
10. 1st Department of Urology, Medical University of Lodz, Lodz, Poland
11. Department of Clinical Chemistry, University of Helsinki, Helsinki, Finland
12. Laboratory Services, Mehiläinen Oy, Helsinki, Finland
13. Mehiläinen Länsi-Pohja Hospital, Kemi, Finland
14. Department of Radiology, Aarhus University Hospital, Aarhus, Denmark
15. Department of Clinical Medicine, Aarhus University, Aarhus, Denmark
16. Department of Molecular Medicine, Aarhus University Hospital, Aarhus, Denmark
17. Department of Pathology, Aarhus University Hospital, Aarhus, Denmark
18. Faculty of Medicine and Health Technology, Tampere University, Tampere, Finland
19. Department of Pathology and Molecular Medicine, Wellington School of Medicine and Health Sciences, University of Otago, Wellington, New Zealand
20. Aquesta Uropathology and University of Queensland, QLD, Brisbane, Australia
21. Department of Surgical Pathology, School of Medicine, Aichi Medical University, Nagoya, Japan
22. Faculty of Science and Technology, University of Stavanger, Stavanger, Norway
23. Department of Oncology and Pathology, Karolinska Institutet, Stockholm, Sweden

### Revision history

| Version and date | Status | Main changes |
|---|---|---|
| 1.0 (2024-07-04) | Initial publication. | Initial publication. |

### 1. AI system design considerations

The AI system development will follow a comprehensive experimental pipeline driven by specific requirements and algorithmic considerations determining the model design and hyperparameter choices. These considerations encompass various aspects of AI design such as the granularity of data annotations, memory constraints, class imbalance, loss functions, training optimisation, and visualisation outputs. Below we list the design choices, which will be systematically evaluated during model development:

- Data preprocessing:
  - Explore different image preprocessing parameters (e.g. hue, saturation, morphological operations like closing size etc.) and convolutional neural network (CNN) algorithms to segment the tissue from the background.
  - Determine the optimal parameters for extracting tiles from whole slide images (WSIs), i.e. tile size, pixel size (2.0 µm, 1.5 µm, 1.0 µm, 0.5 µm), the extent of overlap between adjacent tiles and the minimum amount of tissue required in each tile. These configurations should balance computational efficiency, memory consumption and the level of granularity needed for accurate predictions.
  - Investigate the effect of the chosen tiling parameters, separately, for model training and prediction, considering the different memory consumption constraints during each phase. Investigate if combined tiling parameters affect the performance.
- Data storage:
  - Explore different disk-friendly formats for storing tiles, due to the large number of tiles produced per WSI (Abadi *et al.*, 2016).
- Bias and data quality:
  - Address biases related to the data that could potentially mislead the model during training, e.g. confounding factors on the slides (pen mark annotations from pathologists) or unequal class distributions between slides scanned with different scanning equipment.
- Data augmentation:
  - Apply diverse augmentations to increase robustness to variations in the input data and enhance the model's ability to generalise to unseen samples.
  - Investigate different augmentation techniques such as affine transformations (rotations, vertical or horizontal flipping), noise-simulating augmentations (Gaussian noise, ISO noise, image compression), colour and blur augmentations, and stain augmentations (Faryna, van der Laak and Litjens, 2024).
  - Investigate the effect of test time augmentation (TTA) techniques only, or in addition to the data augmentation during training.
  - Investigate if data augmentations should be conducted on the tile level or the WSI level.

    - Explore how to best utilise different WSIs of the same biopsy slide during model training e.g. sample a random WSI per slide or use all WSIs on each training epoch.
    - Explore the effect of using physical scanner calibration as a data augmentation or normalisation technique (Ji *et al.*, 2022).
- Training data:
    - Sequentially incorporate additional development cohorts as training data.
    - Evaluate the effect of adding more training data for generalising better across different labs and pathology scanners.
- Model design:
    - Investigate learning strategies for handling bags of data where annotations are available at the WSI level rather than the tile level, e.g. self-supervised learning or weakly-supervised learning.
    - Explore design ideas from the top-performing PANDA teams (Bulten *et al.*, 2022).
    - Investigate the effect of encoder architectures and the number of parameters e.g. ResNet18, ResNet34, ResNet50, ResNet101 (He *et al.*, 2016), EfficientNetV2-S, EfficientNetV2-M and EfficientNetV2-L (Tan and Le, 18--24 Jul 2021) or foundation models (Chen *et al.*, 2024; Xu *et al.*, 2024).
    - Investigate aggregation mechanisms for obtaining both WSI level predictions and tile level visualisations (e.g. attention heatmaps).
    - Explore multitask learning for predicting more than one outcome with a single model, e.g. Gleason score (GS) and cancer length.
- Model hyperparameters:
    - Experiment with different conventional learning rates (LR) (e.g. 0.0001, >0.0001, <0.0001) or cyclical LR for balancing learning speed and training stability.
    - Experiment with different optimisers (e.g. Adam, AdamW, SGD) for better convergence (Kingma and Ba, 2014).
    - Experiment with different batch sizes: different number of WSIs and number of tiles per WSI.
- Loss functions:
    - Experiment with different loss functions: regression loss (mean squared error, MSE or mean absolute error, MAE), ordinal loss, cross-entropy loss, Coral loss (Sun and Saenko, 2016) etc.
- Memory optimisation and computational efficiency:
    - Experiment on available hardware on high-performance computing clusters to support larger effective batch sizes without exceeding memory limits.
    - Explore the effect of mixed precision during training and prediction for reduced memory consumption and increased throughput (Micikevicius *et al.*, 2017).

    - Explore the effect of checkpointing gradients (Rumelhart, Hinton and Williams, 1986) for reduced memory footprint at the cost of additional computation time during training.
    - Explore the effect of accumulating gradients over multiple mini-batches before performing the optimiser to obtain larger effective batch sizes without requiring additional memory.
    - Explore employing PyTorch Distributed Data-Parallel (DDP) for multi-node, multi-graphical processing unit (GPU) training to speed up training (Paszke *et al.*, 2017).
- Model output:
    - Investigate different model heads for predicting Gleason patterns, GS, or the International Society of Urological Pathology (ISUP) grade.
    - In the case of predicting GS or patterns using a regression loss function, investigate how to best map those into ordinal variables (Jung *et al.*, 2022; Egevad, Micoli, Delahunt, *et al.*, 2024; Egevad, Micoli, Samaratunga, *et al.*, 2024).
- Early stopping criteria:
    - Experiment with monitoring the validation loss or other metrics (e.g. Cohen's linearly or quadratically weighted kappa, LWK/QWK) and halting training when the metric reaches a plateau.
    - Monitor performance and optimise early stopping parameters, including the minimum and the maximum number of epochs and the patience threshold for terminating training.
- Handle class imbalance:
    - Experiment with a weighted loss function to penalise misclassifications of the minority class more heavily than the majority class.
    - Experiment with sampling to balance class frequencies for each training epoch.
- Model ensembles:
    - Experiment with model ensembles across and within cross-validation folds.
    - Experiment with using different random number generator seeds for each cross-validation fold.
    - Define an optimal number of within-fold and across-fold models for the ensemble.
    - Experiment with varying methods of ensembling e.g. hard-voting and soft-voting.

### 2. Verification of dataset coherence

The datasets undergo rigorous testing to maintain data integrity and accuracy. Below is a list of all tests performed programmatically whenever the dataset is updated. Here, "main database" refers to a spreadsheet containing all the data, where each row represents a single WSI along with its associated pathology information, scanning and digitisation details and data partition. Each WSI, slide (i.e. one glass slide containing one or more tissue cores), “location” (i.e. a set of slides treated as a single diagnostic unit and graded together, sometimes called “part” or “block” but not to be confused with a paraffin block), biopsy (i.e. a single operation during which multiple tissue cores are obtained from the prostate) and patient in the database is represented by a unique identifier (ID), and the data are partitioned into development, tuning, validation or exclusion.

- Consistency in data partitions:
  - All patients assigned to the data partitions ("development", "tuning", "validation", "exclusion") must be present in the main database.
  - All WSIs in the main database should be assigned to a data partition ("development", "tuning", "validation", "exclusion").
  - All WSIs in the "development" partition and none of the WSIs in the other partitions should be assigned to a cross-validation fold.
  - Patients must not overlap between data partitions (i.e. a patient can belong to one partition and one partition only).
  - Partition assignments must be consistent for all WSIs and slides of a patient.
  - Verify that all the patients belonging to special subsets intended for internal validation are assigned to “validation” (ImageBase, perineural invasion (PNI) validation set, PANDA private test sets, morphological subtypes, SUH repeated scanning set, and SUH patients with re-cuts).

- Consistency for variables across patients, biopsies, slides and WSIs:
  - Patient level variables (e.g. age) and slide level variables (e.g. GS) must be consistent across all WSIs representing the same patient or the same slide, respectively. (The requirement for consistent Gleason scoring is relaxed in the case of slides re-graded for PANDA.)
  - Biopsy level variables must be consistent across the WSIs representing the same biopsy procedure (e.g. age at the time of the biopsy, PSA values etc.) since some cohorts include patients who have undergone multiple biopsies at different times.
  - Location level variables must be consistent across the WSIs representing the same “location”. Location IDs must match slide IDs for cohorts with slide level reporting.

- Validation of variable values:
  - Verify that all variables in the database have at least one non-empty entry.
  - Verify that the following variables are non-empty for all WSIs: WSI ID (identifier unique to each dataset entry), ID (unique identifier for each slide), Patient ID

(unique identifier for each patient), Pathology ID (identifier unique to a single biopsy procedure), path and filename of the WSI, and data cohort.

- There must not be any duplicated WSI IDs.
- There must not be any duplicated WSI paths or filenames.
- GS and ISUP grade per slide, location or patient must have valid values i.e. ("0 + 0", "3 + 3", "3 + 4", "4 + 3", "4 + 4", "3 + 5", "5 + 3", "4 + 5", "5 + 4", "5 + 5") and ("0", "1", "2", "3", "4", "5"), respectively, or be empty.
- If both the GS and ISUP grade per slide/location/patient are present, they must follow the ISUP grade definition (e.g. GS 3 + 3 must be ISUP 1).
- At least one grading variable (GS, GS patient, ISUP, ISUP patient) must be populated, except for the morphological subtype cohorts.
- Quantitative variables (e.g. cancer length, biopsy length) must be non-negative or empty.
- Percentual variables (e.g. cancer percentage) must be 0-100 or empty.
- Variables for cribriform cancer and PNI must be True, False, Borderline or empty.
- Boolean variables must be True, False, or empty.

### 3. CONSORT diagrams

The flow charts below represent CONSORT diagrams for all the data cohorts utilised in this study. Subsequent exclusions will be incorporated in revised versions of this protocol if any additional technical issues arise during model development and validation (e.g. file corruption issues encountered during WSI preprocessing or generating tiles from the WSI). Diagrams were created with BioRender.

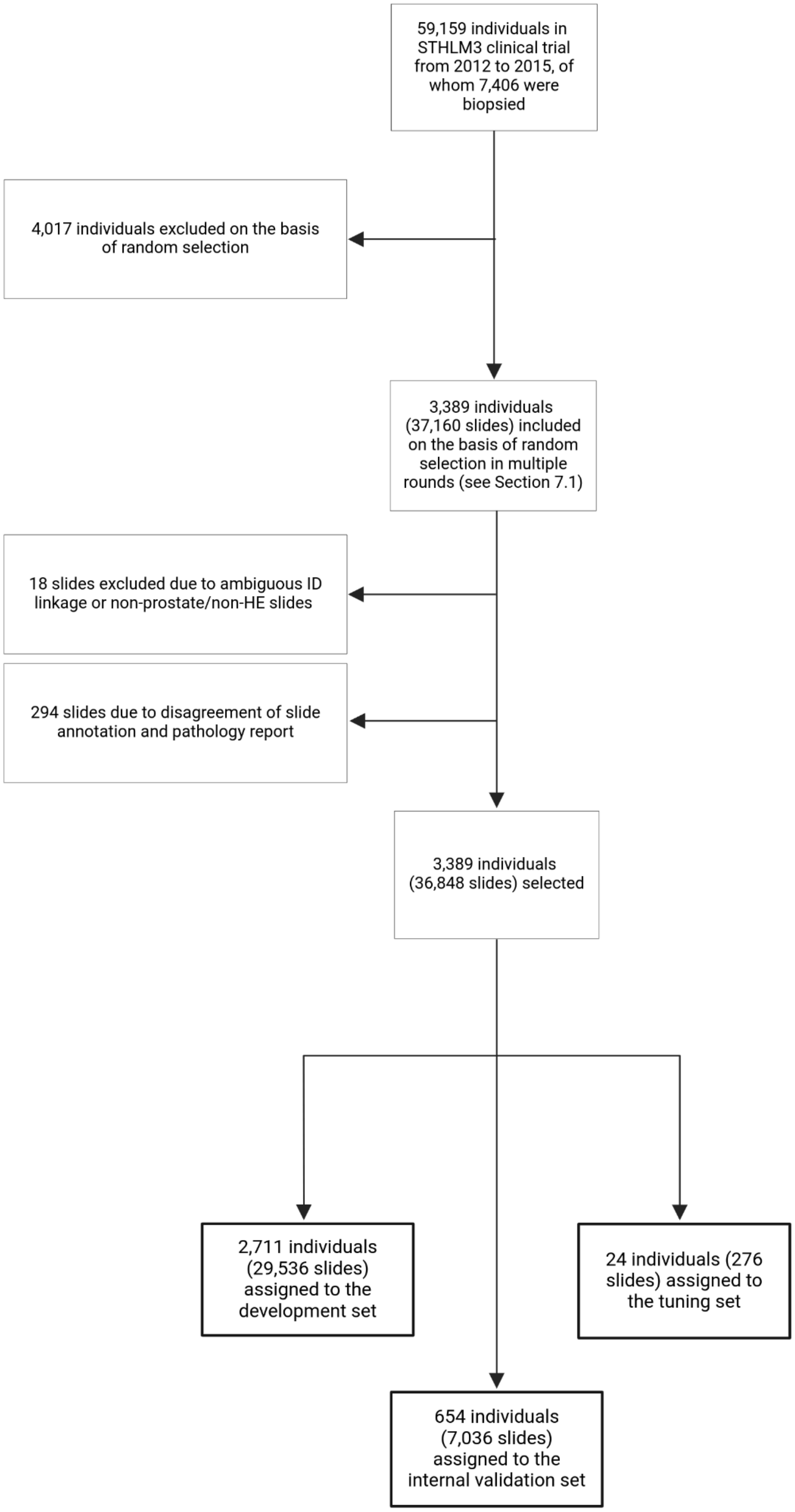

**Figure S1.** CONSORT diagram for the STHLM3 cohort, which is part of the development, tuning and internal validation set.

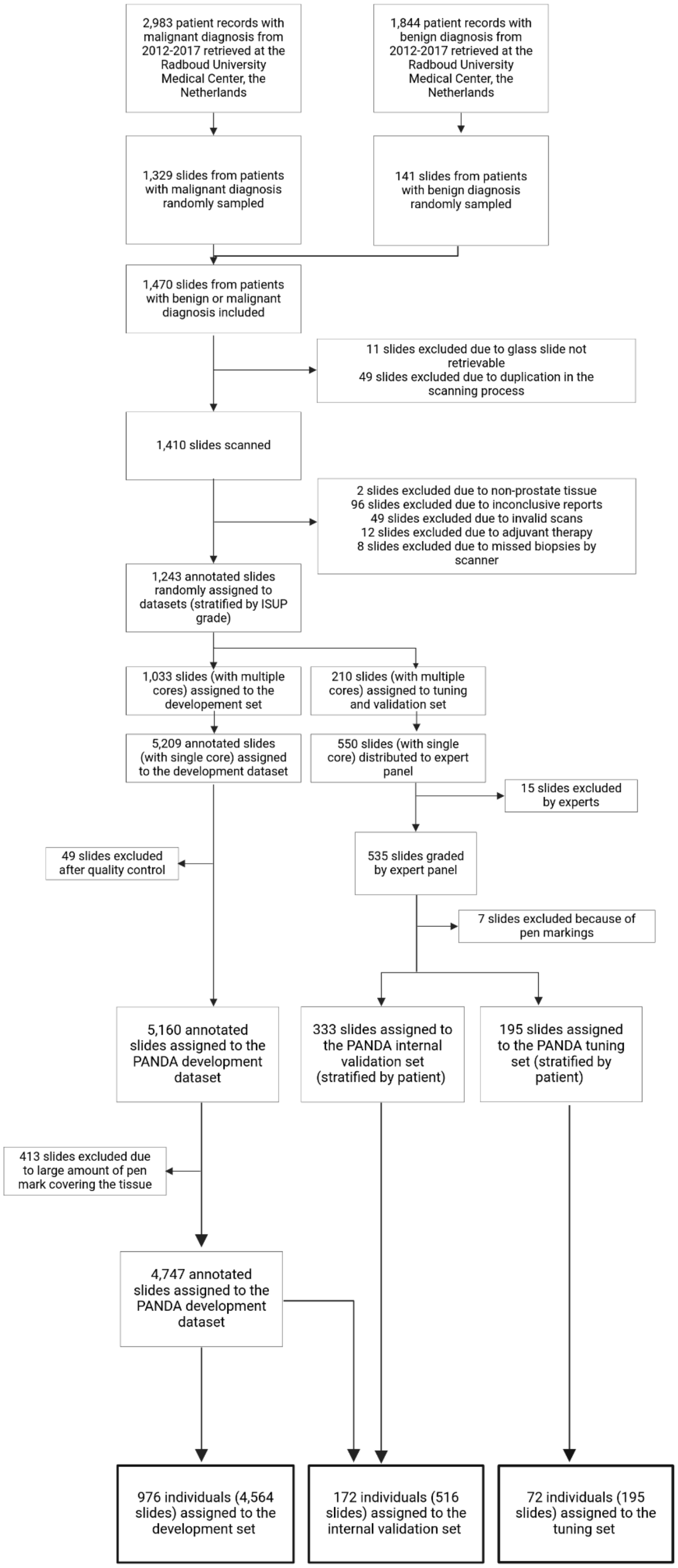

**Figure S2.** CONSORT diagram for the RUMC cohort, which is part of the development, tuning and internal validation set. Adapted from (Bulten *et al.,* 2022).

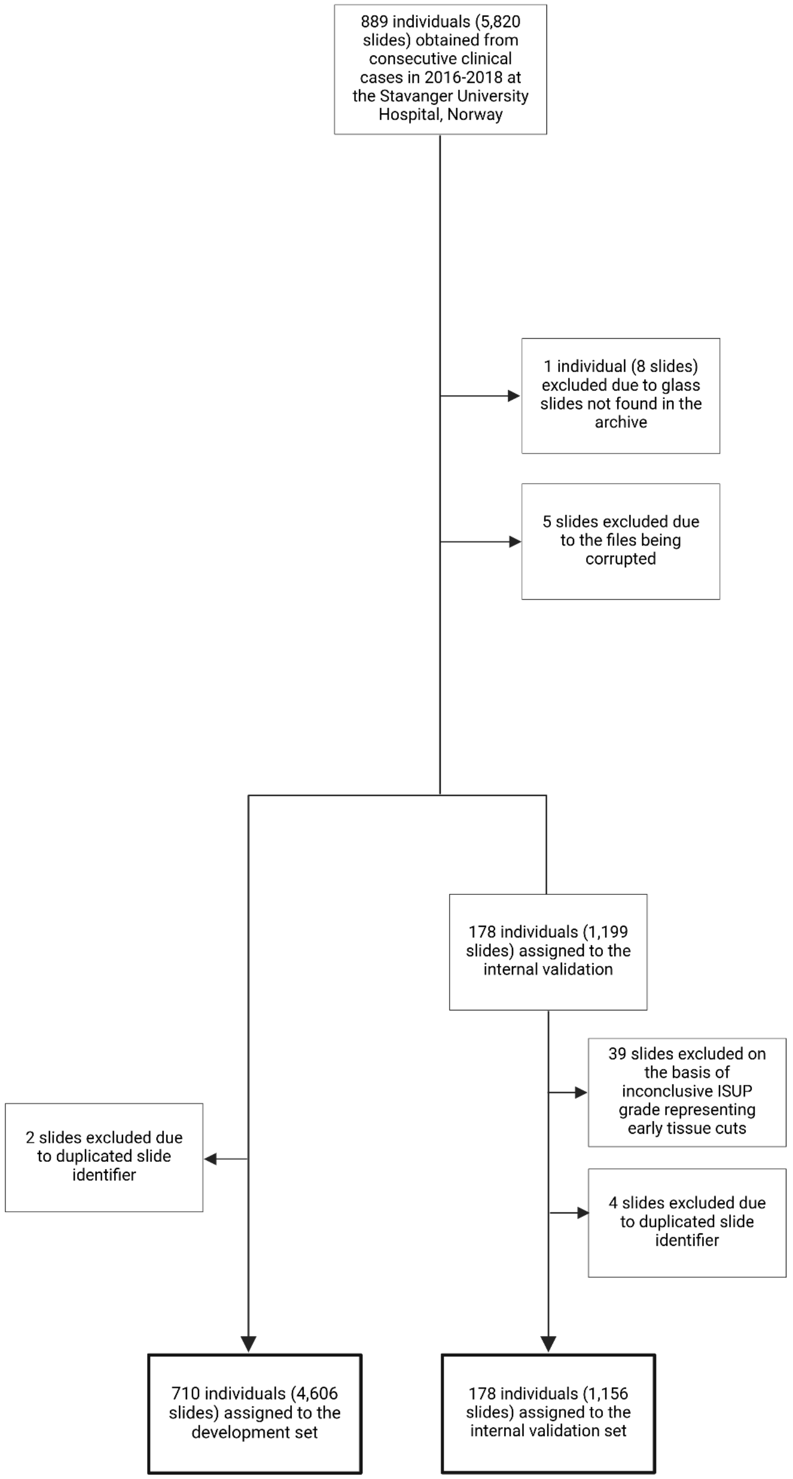

**Figure S3.** CONSORT diagram for the SUH cohort, which is part of the development and internal validation set.

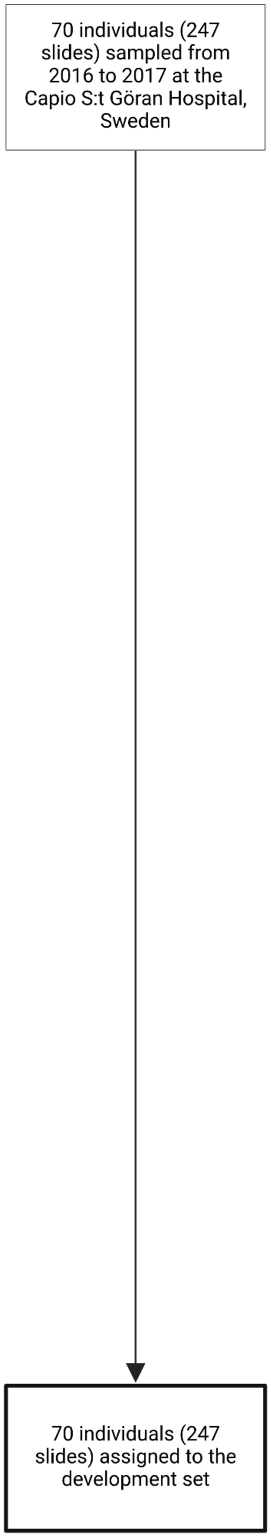

**Figure S4.** CONSORT diagram for the STG cohort, which is part of the development set.

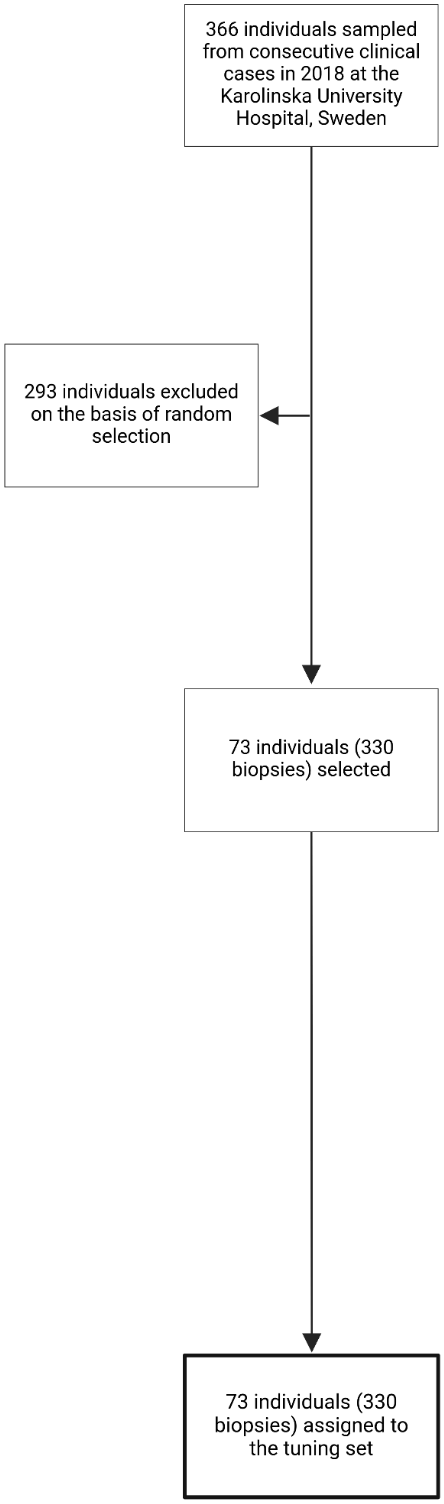

**Figure S5.** CONSORT diagram for the KUH-1 cohort, which is part of the tuning set.

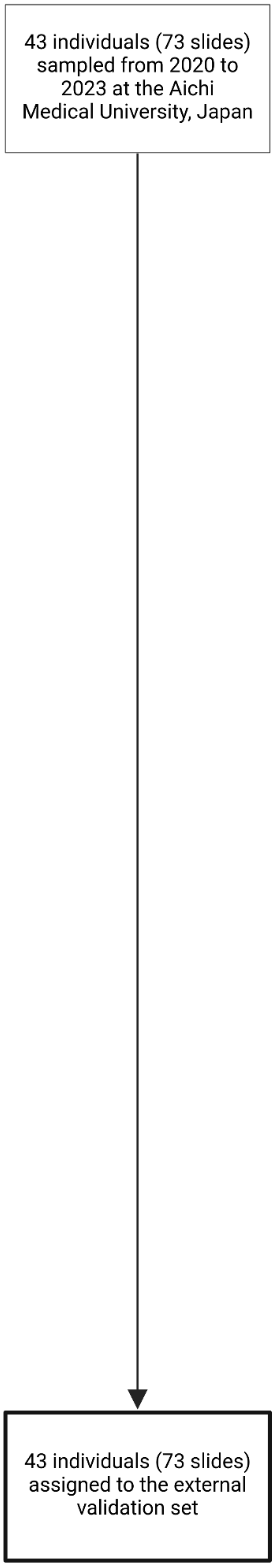

**Figure S6.** CONSORT diagram for the AMU cohort, which is part of the external validation set.

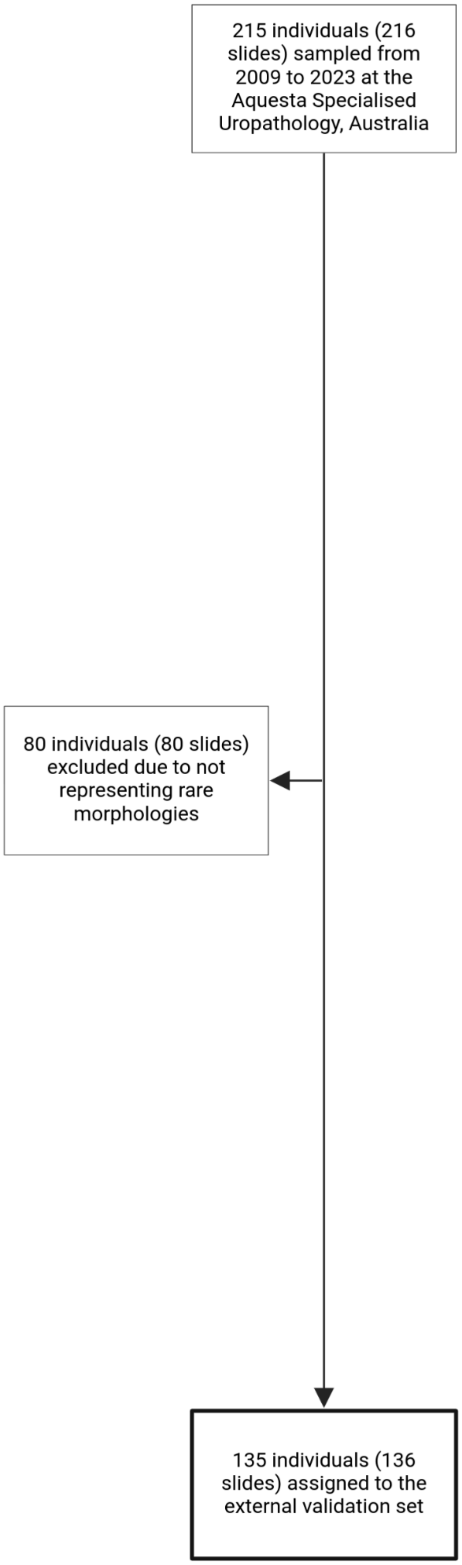

**Figure S7.** CONSORT diagram for the AQ cohort, which is part of the external validation set.

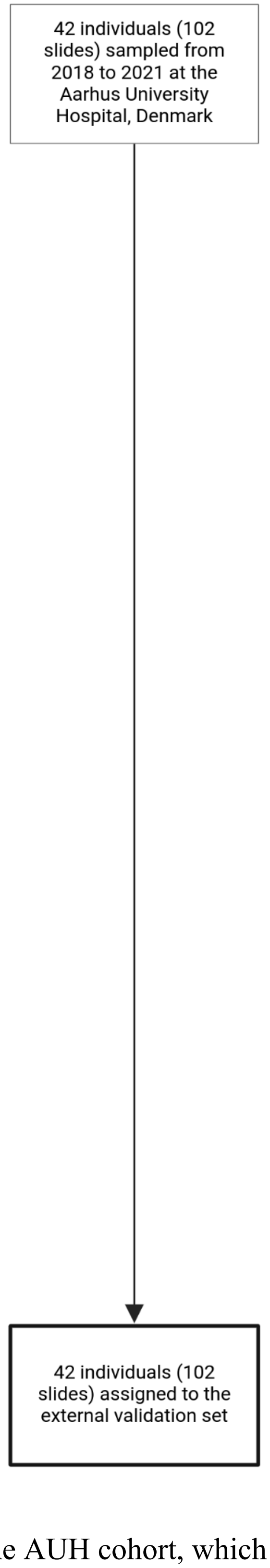

**Figure S8.** CONSORT diagram for the AUH cohort, which is part of the external validation set.

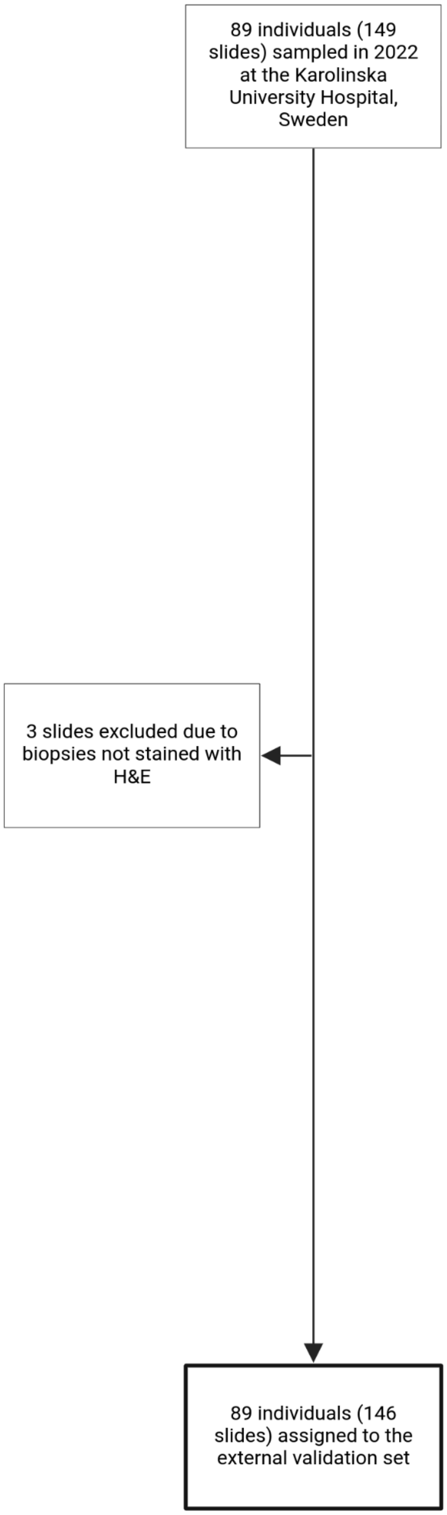

**Figure S9.** CONSORT diagram for the KUH-2 cohort, which is part of the external validation set and represents rare morphologies.

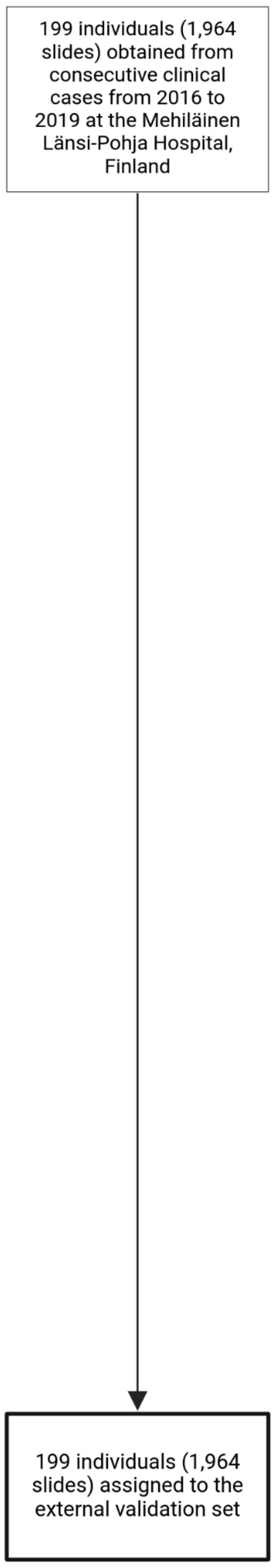

**Figure S10.** CONSORT diagram for the MLP cohort, which is part of the external validation set.

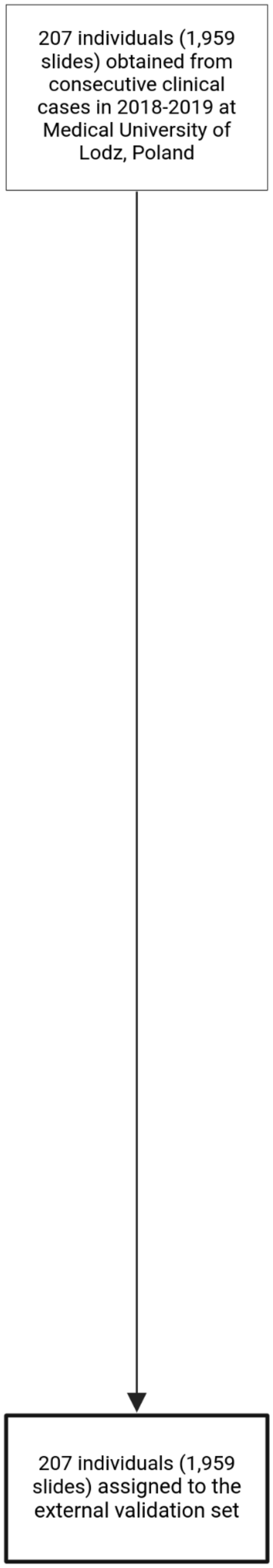

**Figure S11.** CONSORT diagram for the MUL cohort, which is part of the external validation set.

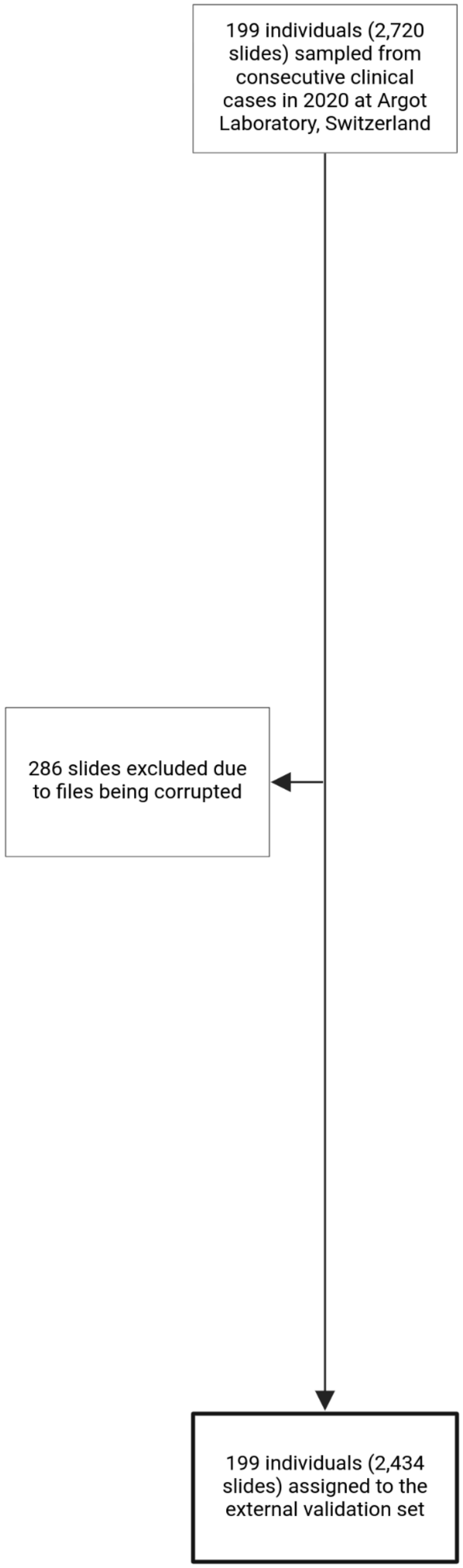

**Figure S12.** CONSORT diagram for the SCH cohort, which is part of the external validation set.

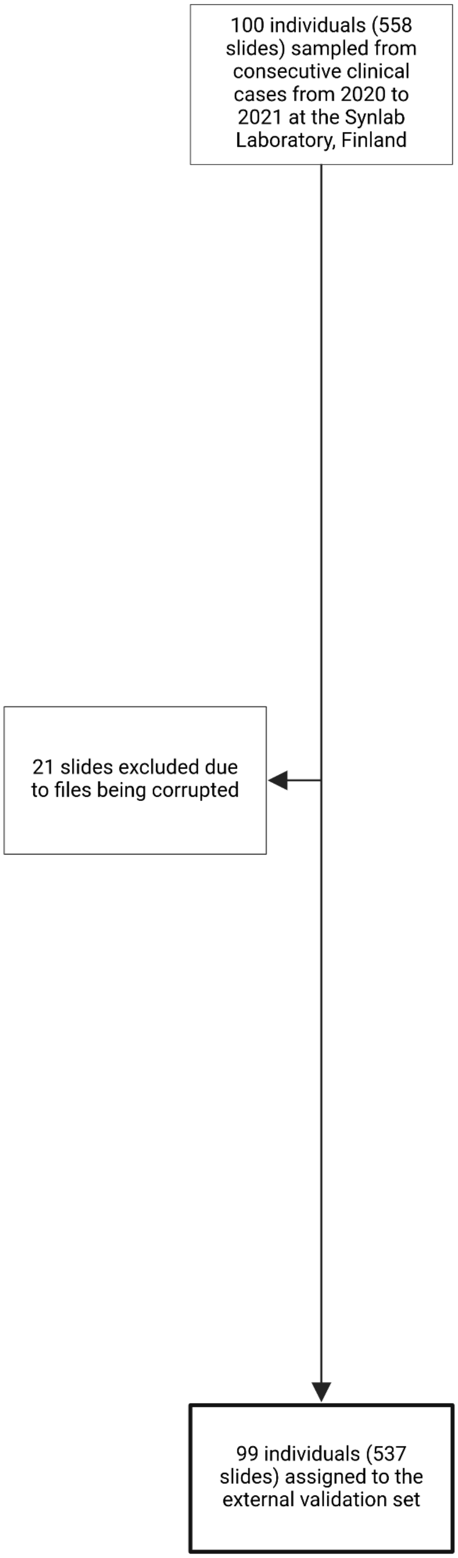

**Figure S13.** CONSORT diagram for the SFI cohort, which is part of the external validation set.

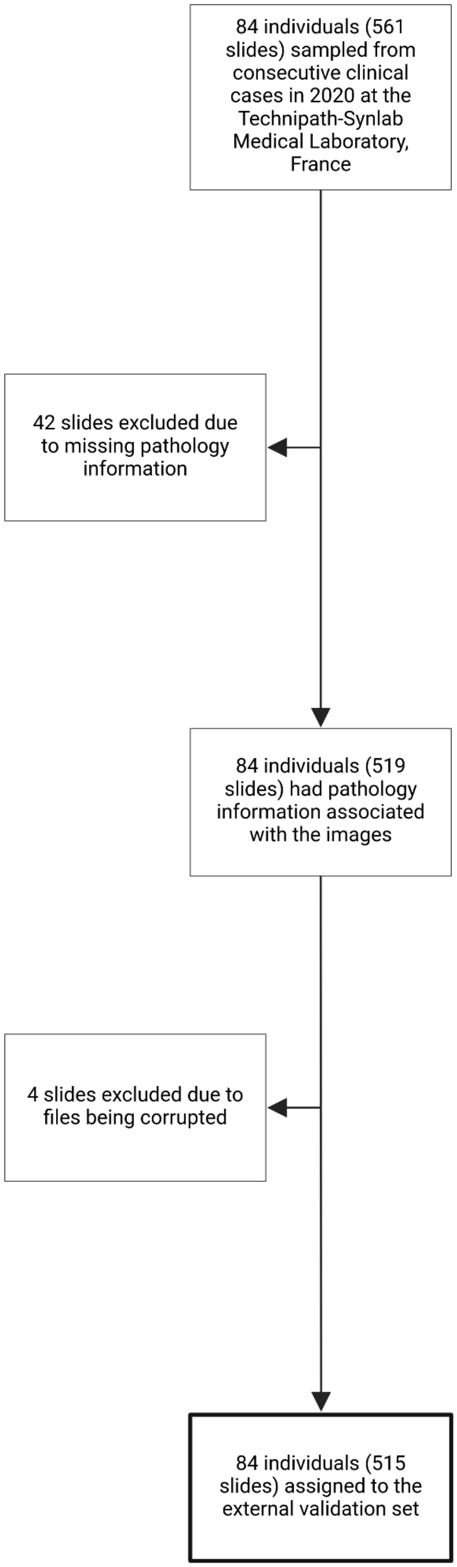

**Figure S14.** CONSORT diagram for the SFR cohort, which is part of the external validation set.

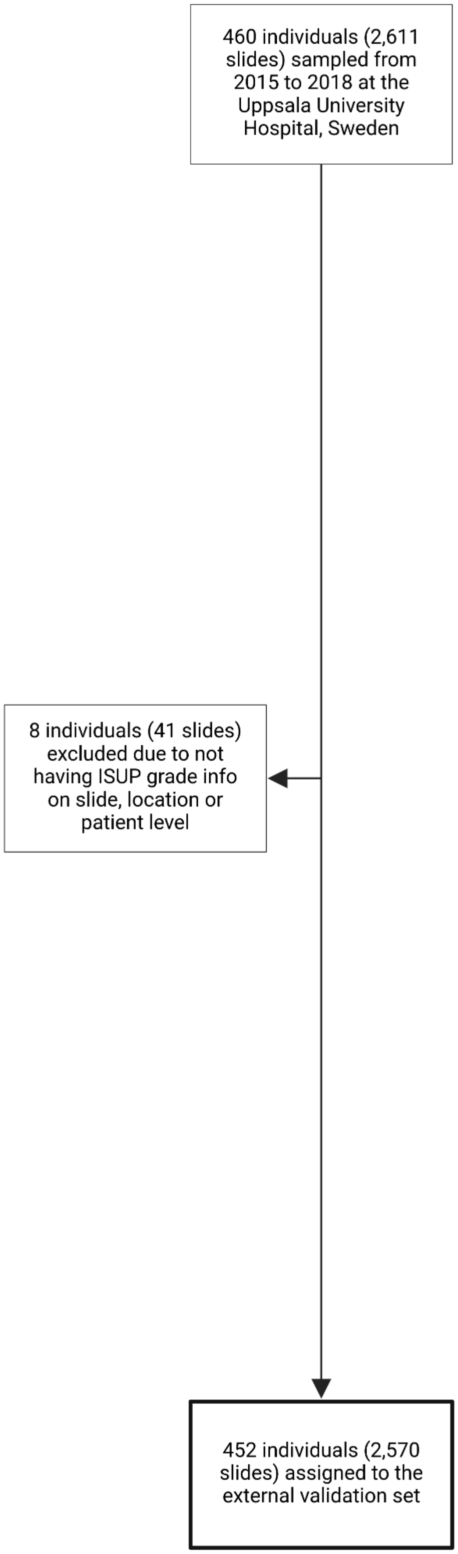

**Figure S15.** CONSORT diagram for the SPROB20 cohort, which is part of the external validation set.

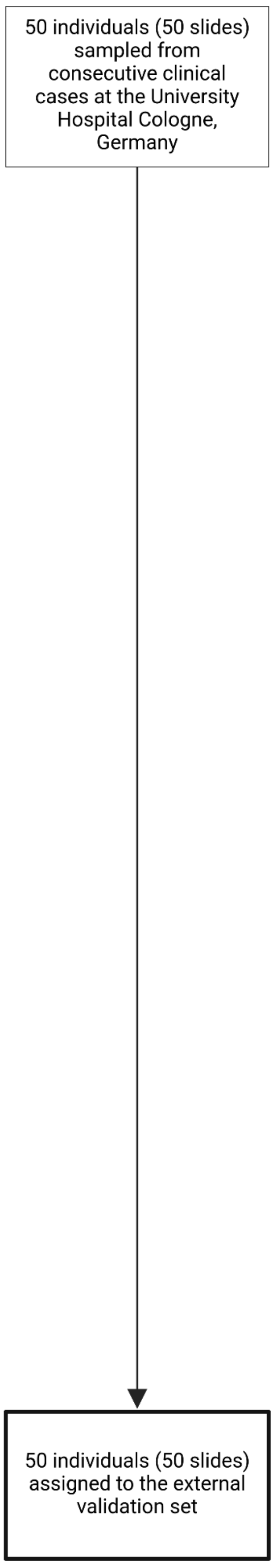

**Figure S16.** CONSORT diagram for the UKK cohort, which is part of the external validation set.

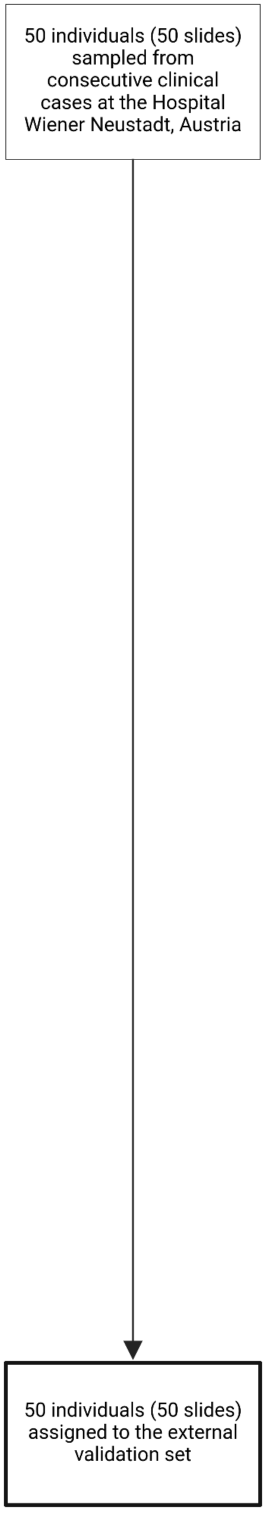

**Figure S17.** CONSORT diagram for the WNS cohort, which is part of the external validation set.
